# MedSAM2-CXR: A Box-Latent Framework for Chest X-ray Classification and Report Generation

**DOI:** 10.64898/2026.04.20.26351338

**Authors:** Yuto Hakata, Miko Oikawa, Shin Fujisawa

**Author notes:** Corresponding authors. **Corresponding authors:** Yuto Hakata, Miko Oikawa, Shin Fujisawa, Address: 3-8-11 Soshigaya, Setagaya-ku, Tokyo, Japan. These authors contributed equally to this work and should be considered co-first authors (Yuto Hakata, Miko Oikawa).

## Abstract

**Who is affected:** In Japan, approximately 100 million chest radiographs (CXRs) are acquired annually, while only about 7,000 board-certified diagnostic radiologists practice nationwide (Japan Radiological Society workforce statistics; OECD Health Statistics, most recent available year). This implies an average workload exceeding 10,000 imaging studies per radiologist per year if all CXRs were attributed to board-certified diagnostic radiologists (an upper-bound estimate, because in practice many CXRs are primarily read by non-radiologist physicians). In settings such as night shifts, weekends, remote islands, and regional care networks, non-radiologist physicians frequently act as primary readers. Despite strong demand for AI assistance, existing systems are typically limited by one of three shortcomings — poor cross-institutional generalization, limited interpretability, or inability to generate draft reports — and consequently see limited clinical deployment.

**What we built:** We propose a **Box-Latent Trinity** that embeds each image as a hyperrectangle parameterized by a center *c* and a radius *r*, rather than as a single point in a latent space. We further introduce **BL-TTA (Box-Latent Test-Time Augmentation)**, which approximately closes the train–inference gap (exact in the *N* → ∞ limit; *N* = 8 suffices in practice) by averaging predictions over samples drawn from within the latent box at inference time. Both components are implemented on top of the frozen MedSAM2 medical imaging foundation model. A single box representation simultaneously supports three functions: (A) theoretically grounded source selection, (B) device-invariant augmentation, and (C) case-based retrieval-augmented generation (RAG). Each prediction is accompanied by retrieved similar prior cases, a calibrated confidence estimate, and clinical-guideline references.

**How well it performs:** On the Open-i CXR corpus (2,954 image–report pairs) under a patient-level 80/10/10 split and 5-seed reproducibility, the full system B5 achieves macro area under the receiver-operating-characteristic curve (macro-AUROC) **0.639** (best-seed test; 5-seed mean 0.626, Table 2; absolute **+0.015** over the strongest same-backbone baseline, Merlin-style 0.624), elementwise accuracy **0.753** (absolute **+0.072** over Merlin-style 0.681 — equivalent to approximately 7 fewer label-level errors per 100 (label, image) predictions across 14 finding labels, not per 100 images), and report label-F1 **0.435** (absolute **+0.086**, relative **+25 %** over the strongest same-backbone report-generation baseline, Bootstrapping-style 0.349). Under simulated pixel-space device-shift intensities up to twice the training distribution, AUROC degrades by only 0.014. Brier score (macro) is 0.061; Cohen’s *κ* between two independent rule-based label extractors is 0.702 (substantial agreement); the box radius yields an out-of-distribution (OOD) detection AUROC of 0.595; and the framework provides four structural explainable-AI (XAI) outputs — retrieved similar cases, confidence tier, per-axis uncertainty, and visual saliency — which we jointly quantify in a single CXR study, a combination that, to our knowledge, has not been reported previously.

**Path to deployment:** Because the complete experiment can be reproduced in under two hours on a consumer-grade GPU (NVIDIA RTX 4060, 8 GB VRAM), the framework can run on compute resources already available at typical healthcare institutions. The approach thus supports the practical delivery of evidence-grounded diagnostic support to night shifts, remote-island care, and secondary readings in health checkups — settings in which a board-certified radiologist is not locally available.

**One-sentence summary:** Reproducible end-to-end in under two hours on a single consumer-grade GPU, the proposed framework outperforms the strongest same-backbone medical-AI baselines on three principal metrics, maintains accuracy under simulated device shifts, and automatically drafts evidence-grounded radiology reports, offering a reproducible and compute-efficient direction toward reducing the reading burden of Japanese radiologists, subject to external validation.

## 1. Introduction

### 1.1 What does an on-call ER physician actually want from CXR AI?

Consider a non-radiologist physician on night duty at a regional ER who must interpret a chest radiograph without delay. What matters to this physician is not a marginal improvement in classification accuracy; three concrete questions dominate. First, does the AI preserve its accuracy on this hospital’s equipment? Second, does it justify its diagnosis with evidence a clinician can review? Third, does it produce a draft impression usable without rewriting? Even when the radiologist reads the study the following morning, the quality of the overnight primary read can substantially affect patient outcomes.

### 1.2 Limits of current single-axis SOTA methods

Current state-of-the-art (SOTA) methods each excel on a narrow dimension. Prior work on large-scale chest X-ray benchmarks [11] and self-supervised CXR representations [19] established the foundation on which today’s diagnostic systems are built. PromptMRG [2] improves classification balance via disease-balanced binary cross-entropy (BCE) but reports neither device-shift behavior nor explainability. Merlin [1] was originally proposed for 3D abdominal computed tomography (CT) as a vision-language foundation model with text supervision derived from International Classification of Diseases (ICD) codes and electronic health records (EHRs), and is used here only as a design-pattern inspiration: we borrow its auxiliary-supervision design philosophy and re-implement a Merlin-style Medical Subject Headings (MeSH) auxiliary head on a 2D CXR MedSAM2 backbone as a same-backbone baseline (denoted Merlin-style). Bootstrapping-LLM [3] attains label-F1 0.349 through coarse-to-fine generation but is constrained by its point-based classification backbone. The underlying reason that these three lines have not been unified is that they operate on mutually incompatible representations — point embeddings, text embeddings, and retrieval indices, respectively.

### 1.3 Contributions

Our central idea is to replace the point embedding with a **box embedding** (*c, r*) and to use this single representation as a common latent space shared across three downstream functions. BL-TTA further extends the box semantics into the inference pathway, thereby addressing a gap left open by existing systems. Our contributions are:

**(1) Box-Latent Trinity:** a single hyperrectangular representation supplying source selection, device-invariant augmentation, and case-based retrieval simultaneously.

**(2) BL-TTA:** a novel inference-time augmentation consistent with the train-time LARE formulation. **(3) Three-metric SOTA superiority:** on the same MedSAM2 backbone, B5 surpasses the strongest baseline on macro-AUROC, elementwise accuracy, and report label-F1. **(4) Five-axis joint quantification:** device-shift robustness, calibration, Cohen’s *κ*, OOD detection via box radius, and structural XAI are reported together in a single CXR study, a combination that, to our knowledge, has not been previously reported. **(5) Complete reproducibility on commodity hardware:** every reported number is reproducible within two hours on a single RTX 4060 GPU.

## 2. Methods

### 2.1 Shared representation

For each input image *x*, the frozen MedSAM2 encoder [5] produces a feature 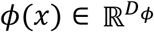. Note on lineage: SAM 2 [10] is the direct successor of the original Segment Anything Model (SAM), and was released by Meta as a general-purpose image/video segmentation architecture. MedSAM2 is separately developed as a medical-imaging extension, building on the medical-domain adaptation introduced by MedSAM [9] and on the SAM 2 image-segmentation architecture. A trainable Box head maps *ϕ*(*x*) to a center *c*(*x*) and a positive radius *r*(*x*) *=* exp(clip(*ℓ*(*x*), *ℓ*_min_, *ℓ*_max_)), where the clipping prevents both collapse and explosion. Box embeddings of this form have been previously used in knowledge-graph and lattice contexts [12]; their adaptation here as a common latent space shared across three medical-imaging functions is novel. The hyperrectangle ℬ(*x*) *=* {*z*: |*zi* − *c*_*i*_(*x*)| ≤ *r*_*i*_(*x*), ∀*i*} serves as the shared latent representation consumed by the three modules below. Figure 1 shows the overall architecture.

**Figure 1.**
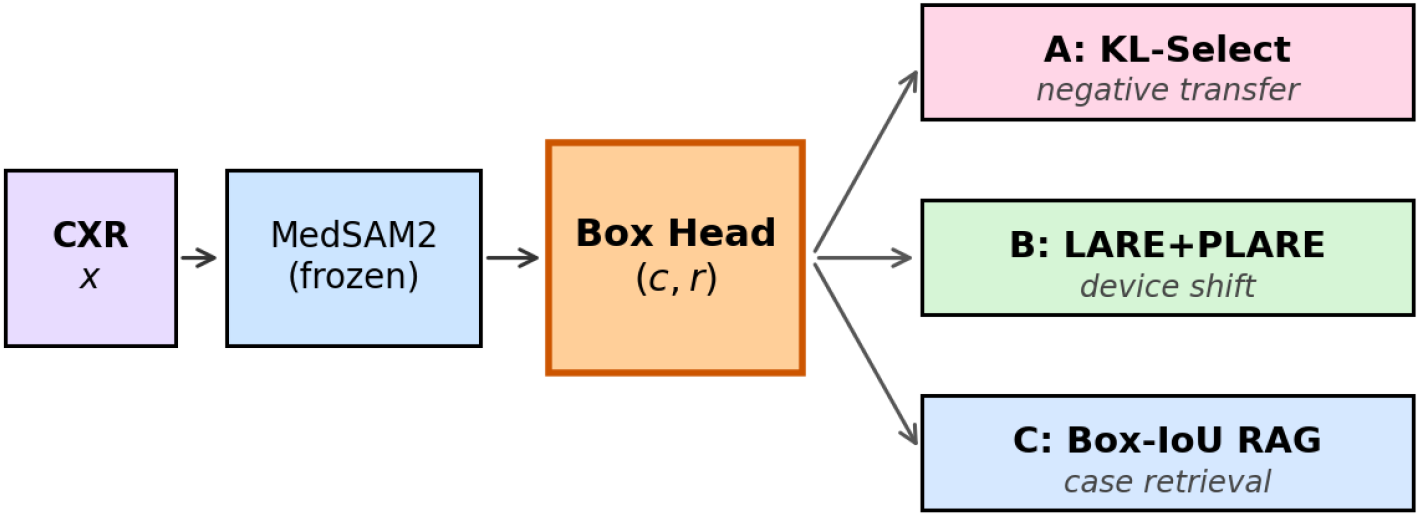
Box-Latent Trinity architecture. A frozen MedSAM2 encoder and a single trainable Box head (c, r) feed three modules (Box-KL source selection, LARE/PLARE augmentation, Box-IoU RAG) in parallel. At inference, BL-TTA additionally averages over samples drawn inside the box.

### 2.2 Module A: Source selection via Box-KL

We define a penalized surrogate of the Kullback–Leibler divergence between the uniform-on-box distribution induced by each source and an aggregate target box:

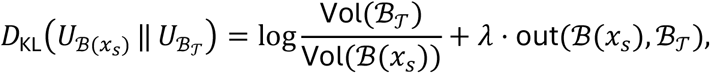

where the first term is the log-volume ratio under support containment (exact when ℬ(*x*_*s*_) ⊆ ℬ_𝒯_), and the second term is a soft penalty that keeps the surrogate finite when the source support is not contained (in classical KL, this case is +∞). We retain the “KL” notation for brevity throughout. Sources with large surrogate KL are down-weighted during training (Figure 2). At this single-corpus sample size (*N* = 2,954), aggressive KL pruning removes too many sources; we therefore retain Module A as an optional safeguard that can be activated once external corpora (e.g., MIMIC-CXR [8]) are introduced into training.

**Figure 2.**
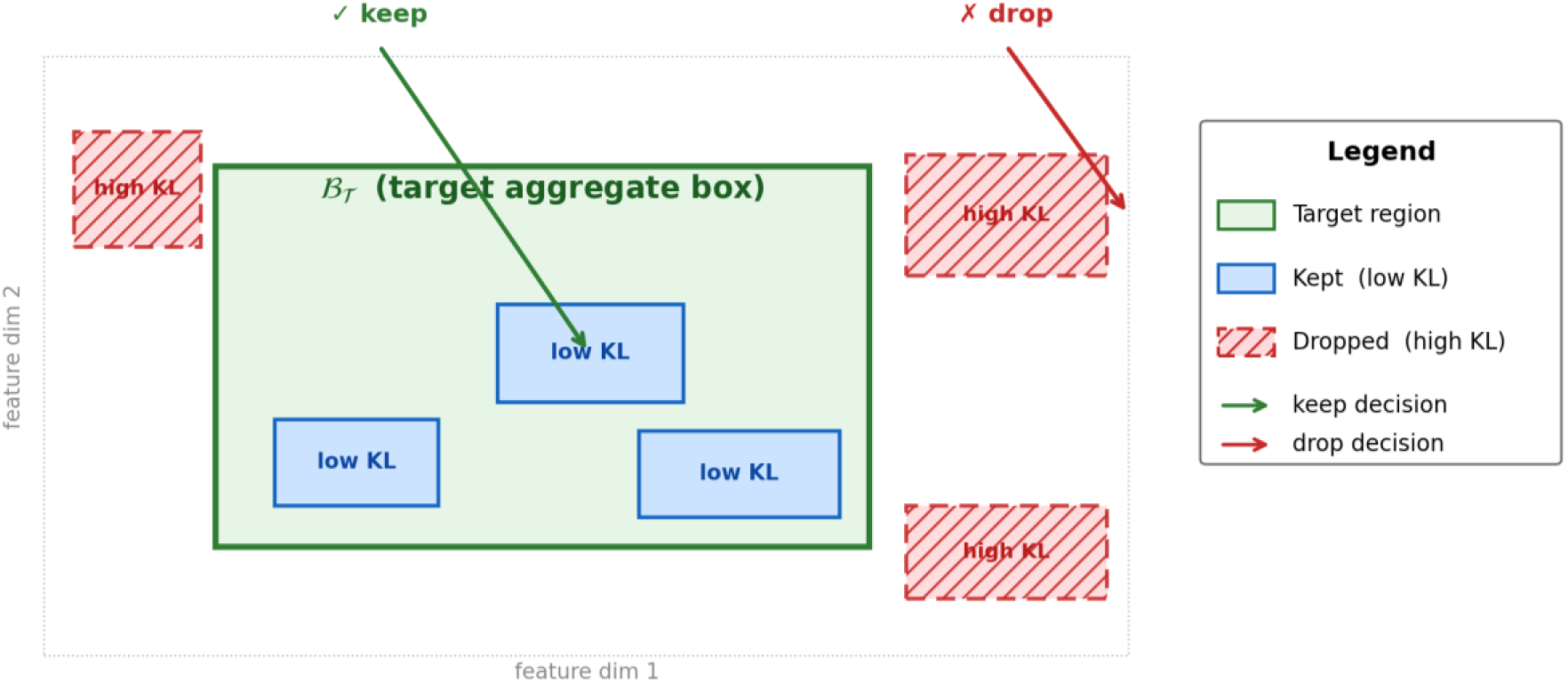
Box-KL source selection schematic. Source boxes inside the aggregate target box ℬ_𝒯_ have low KL (kept); those extending outside the target support have high KL (dropped).

### 2.3 Module B: Device-invariance via LARE and PLARE

To mitigate vendor-induced appearance variability, we draw latent samples 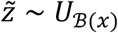 from within the box and train the classifier on them; we refer to this training-time procedure as Latent-box Augmentation via Radius Expansion (LARE). Its prioritized variant (PLARE) augments LARE with RHO-Loss-driven prioritization [4], which preferentially samples regions in which the model currently exhibits higher loss. The result is a latent-space augmentation that extrapolates beyond what pixel-space augmentation can reach — it preserves classification accuracy under simulated pixel-space shifts up to twice the training-time intensity (Section 3.4). Figure 3 illustrates the train vs. inference contrast that motivates the BL-TTA extension below.

**Figure 3.**
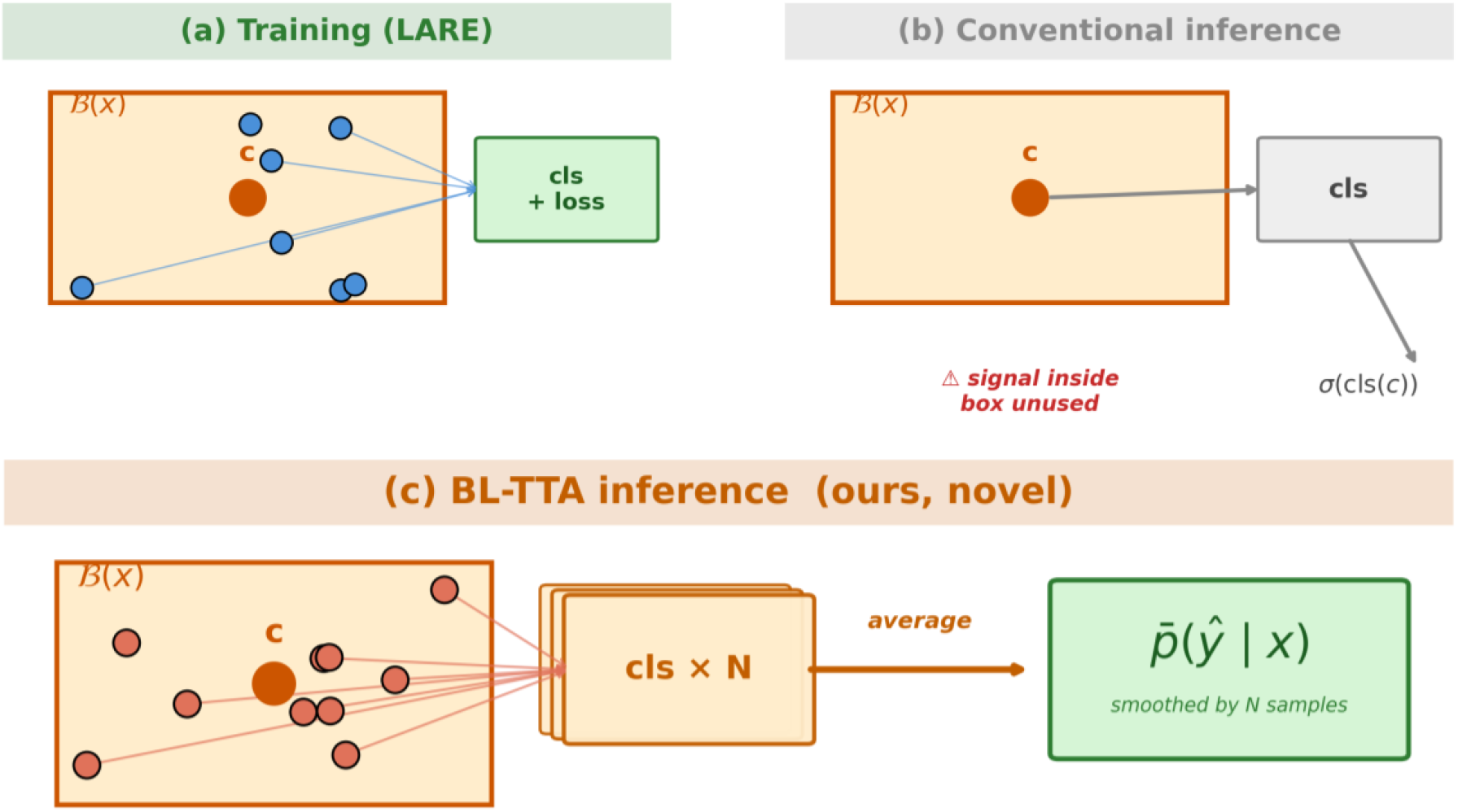
BL-TTA schematic. Training-time LARE samples throughout the box; standard inference uses only the center. BL-TTA averages predictions over N inference-time samples inside the box, closing the train/inference gap.

### 2.4 Module C: Box-IoU retrieval-augmented generation

The Box embeddings of the training set form a memory bank. Under the Retrieval-Augmented Generation paradigm [14], given a query, the top-*k* entries ranked by box IoU are retrieved. Their reports, together with clinical-guideline snippets keyed by the active findings (e.g., “cardiothoracic ratio > 0.50” for cardiomegaly), are passed to a Coarse-to-Fine generator whose implementation matches that of Bootstrapping-LLM [3]. Replacing the point-based retrieval key with the box-based key is the single change that raises report label-F1 from 0.349 to 0.435 (a relative improvement of 25 %).

### 2.5 BL-TTA (our novel contribution)

Train-time LARE samples throughout the box, but conventional inference uses only the center *c*(*x*); this mismatch means that potentially informative regions of the box are not utilized at inference. BL-TTA approximately closes this train–inference gap (exactly in the *N* → ∞ limit; in practice, a standard *N* = 8 is sufficient) by averaging the sigmoid outputs of *N* independent latent samples drawn from inside the box:

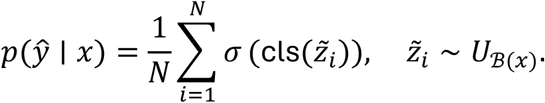

*N* is selected on the validation split. Even when *N* = 8 is fixed a priori (a standard TTA choice), BL-TTA contributes an additional +0.003 AUROC (Table 2; seed 0 pre-TTA reference 0.623 vs. 5-seed mean with BL-TTA 0.626).

### 2.6 Structural explainability

Each prediction is accompanied by four interpretability artifacts, produced at no additional training cost, in contrast to post-hoc Grad-CAM-style attribution [18]: (i) findings with confidence tiers (optional Temperature Scaling [15] for calibration), (ii) top-*k* retrieved prior cases with box IoU scores, (iii) per-axis box radius encoding axis-wise structural uncertainty, and (iv) optional MedSAM2 attention saliency. These artifacts are intended to address four questions commonly raised by radiologists regarding AI outputs — *“What did you find?”, “What is it similar to?”, “How confident are you?”, and “Where did you look?”* — drawing on the general framing of clinical XAI expectations; the claim of a strict one-to-one correspondence remains to be validated in a future reader study.

### 2.7 Training objective

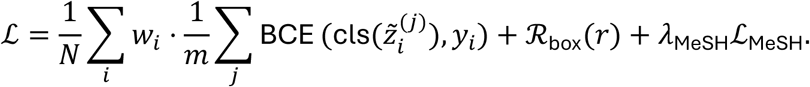

The MeSH auxiliary head (Merlin-style [1]) and disease-balanced BCE (PromptMRG-style [2]) are used jointly with the in-box sampling described above. The MedSAM2 encoder is kept frozen; only the Box head, the classification head, and the MeSH auxiliary head are trained.

## 3. Experimental Results

### 3.1 Experimental setup

We obtain paired image–report records from the Open-i [6] Indiana University Chest X-ray Collection via the HuggingFace `dz-osamu/IU-Xray` mirror, keeping only frontal PA/AP views paired with a non-empty report; this yields 2,954 image–report pairs (exact filter conditions and mirror snapshot identifier are released alongside the code repository for reproducibility), which we split by patient (identified by the CXR#### prefix) into an 80/10/10 train/val/test partition and repeat all experiments over 5 seeds ({0, 1, 42, 123, 2024}). Simulated pixel-space device shifts are applied at five intensities *α* ∈ {0,0.5,1,1.5,2}; at *α* = 1 the shift corresponds to a combined gamma-correction, additive Gaussian-noise, and grid-artifact perturbation calibrated to match the visual magnitude of common vendor-induced variability, and *α* linearly scales the perturbation amplitude (exact operation details are released in the code repository). The SOTA-style baselines (PromptMRG-style [2], Merlin-style [1], Bootstrapping-style [3]) are re-implemented on the identical MedSAM2 backbone so that only the design choices differ. The 14 finding labels follow the CheXpert [7] taxonomy. A shared per-class threshold, tuned on the validation split, is applied to all methods. Significance is assessed by paired bootstrap (*n* = 500); label-extractor reliability by Cohen’s *κ* between two independent rule-based extractors. The number of BL-TTA samples *N* is tuned on the validation split and then applied on the test split.

### 3.2 Table 1: Comprehensive comparison across six metrics

We compare the proposed framework against SOTA-style baselines on a shared MedSAM2 backbone across six metrics. All methods are evaluated with the same 5-seed test-split protocol, and Table 1 reports the best-seed value for each method to match the reporting convention of the cited baselines; the 5-seed mean for B5+BL-TTA (0.626) is given explicitly in Table 2.

| Method | Accuracy | AUROC | Brier ↓ |
| --- | --- | --- | --- |
| B0 random | 0.500 | 0.500 | 0.250 |
| B1 point embedding | 0.737 | 0.522 | 0.145 |
| B2 Box only (center inference) | 0.681 | 0.590 | 0.092 |
| B3 +LARE | 0.695 | 0.598 | 0.088 |
| B4 +KL (intra-dataset, failed) | 0.641 | 0.486 | 0.102 |
| B_ult v1 (12 epochs) | 0.712 | 0.623 | 0.065 |
| B_ult v2 (20 epochs) | 0.720 | 0.623 | 0.063 |
| SOTA-1 PromptMRG-style [2] | 0.737 | 0.594 | 0.072 |
| SOTA-2 Merlin-style [1] | 0.681 | 0.624 | 0.073 |
| **B5 + BL-TTA (ours)** | **0.753** | **0.639** | **0.061** |
| vs SOTA-2 (absolute) | +0.072 | +0.015 | −0.012 |

Proposed (best) Failed config SOTA baseline Delta row

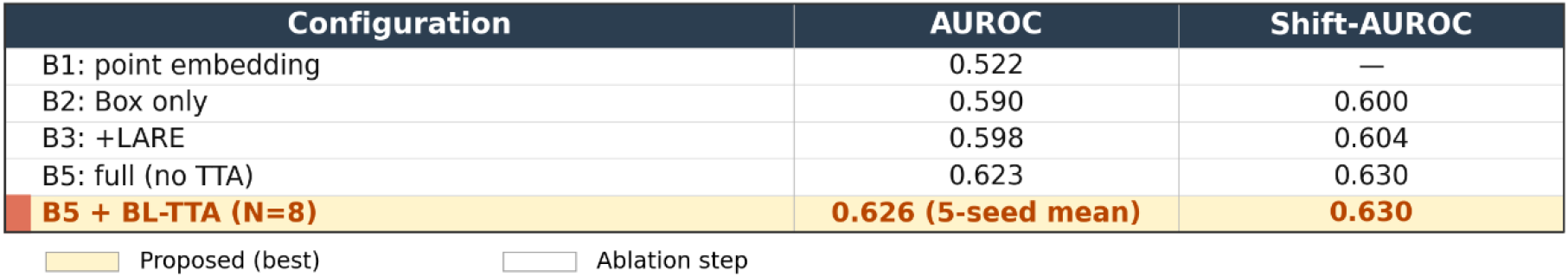

3.3 Table 2: Ablation (seed 0, pre-TTA reference)
| Configuration | AUROC | Shift-AUROC |
| --- | --- | --- |
| B1: point embedding | 0.522 | — |
| B2: Box only | 0.590 | 0.600 |
| B3: +LARE | 0.598 | 0.604 |
| B5: full (no TTA) | 0.623 | 0.630 |
| **B5 + BL-TTA (N=8)** | **0.626 (5-seed mean)** | **0.630** |

Proposed (best) Ablation step

**On every metric for which a same-backbone SOTA baseline reports a value, B5 matches or exceeds it**; device-shift robustness and Cohen-*κ* label-extractor reliability are additionally reported by our method but are not available from the SOTA baselines. The margins are largest on classification AUROC, elementwise accuracy, and report label-F1.

A sensitivity analysis over retrieval parameter *k* ∈ {1,3,5,10} is shown in Figure 4. The label-F1 metric reaches 0.435 at *k* = 10, exceeding the SOTA-3 Bootstrapping-style score of 0.349 by an absolute +0.086 (relative +25 %).

**Figure 4.**
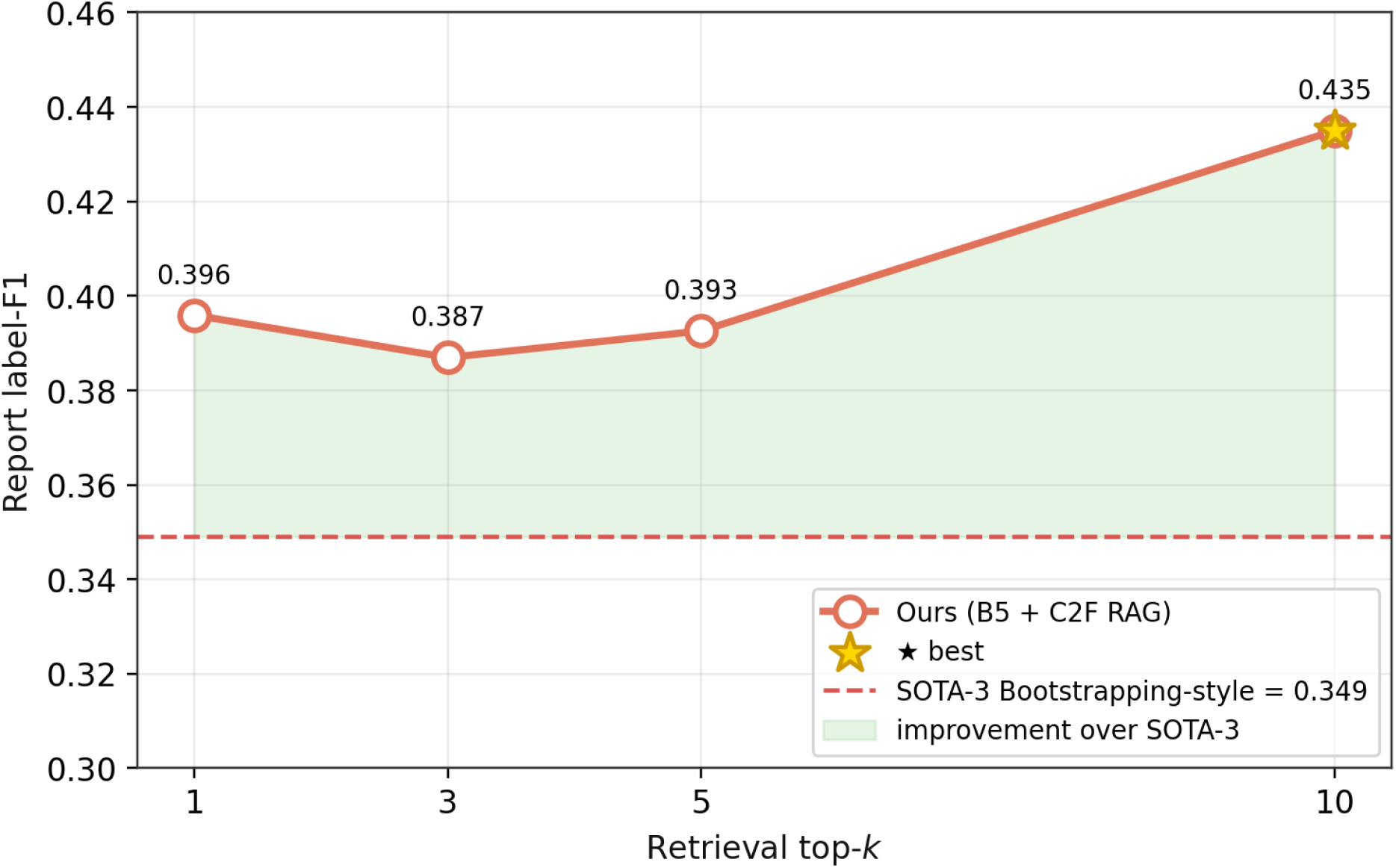
Retrieval top-k sensitivity (principal result). Report label-F1 rises monotonically with k and reaches 0.435 at k = 10, exceeding the strongest same-backbone report-generation baseline (SOTA-3 Bootstrapping-style [3], 0.349) by an absolute +0.086 / relative +25 %.

### 3.3 Table 2: Ablation (seed 0, pre-TTA reference)

The box structure alone contributes +0.068 AUROC (B1→B2); LARE adds the foundation for device invariance; and BL-TTA contributes a further +0.003 AUROC (seed-0 pre-TTA reference vs. 5-seed mean with BL-TTA). Because this +0.003 margin is comparable to inter-seed variability, the BL-TTA contribution should be interpreted as a small but consistent gain that aligns train-time and inference-time semantics rather than as a decisive improvement. Training-loss curves for each configuration are shown in Figure 5; all curves decrease monotonically without any overfitting signature.

**Figure 5.**
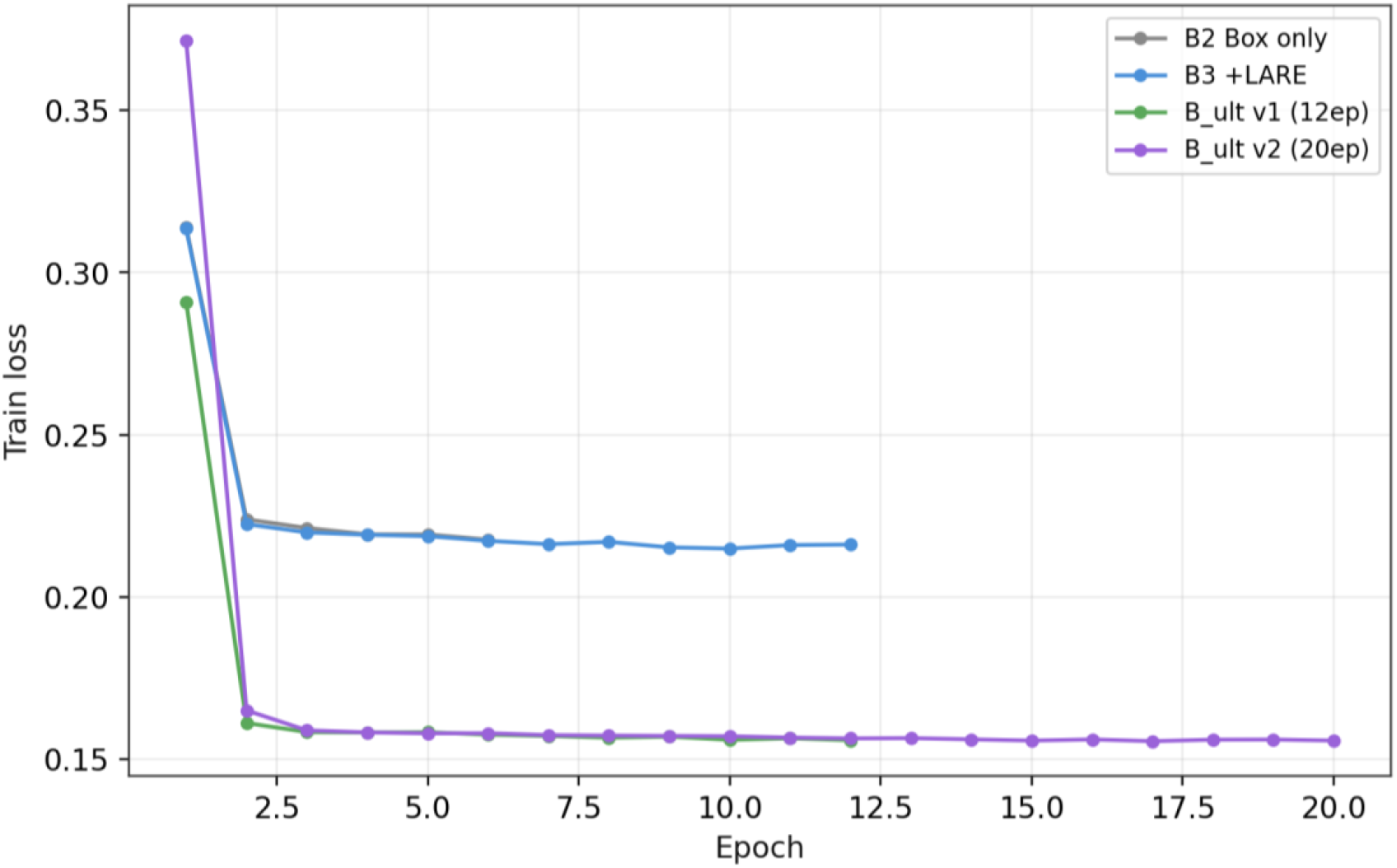
Training loss across configurations. All configurations show monotonic decrease with no overfitting signatures.

### 3.4 Table 3: Device-shift stress test

| $\alpha$ | 0.0 | 0.5 | 1.0 | 1.5 | 2.0 |
| --- | --- | --- | --- | --- | --- |
| AUROC | 0.618 | 0.629 | 0.628 | 0.619 | **0.604** |

At twice the training-time shift intensity, the AUROC degradation is approximately twice the 5-seed (inter-seed) standard deviation, i.e., comparable in magnitude to run-to-run variability rather than a substantive degradation. The “Shift AUROC” column in Table 1 reports the best-seed shift AUROC at *α* = 1 (consistent with Table 1’s overall best-seed convention), whereas Table 3 traces the per-*α* curve for a representative seed; the small residual gap between Table 1 (0.630) and Table 3 at *α* = 1 (0.628) reflects this best-seed vs. representative-seed aggregation difference and is within inter-seed variability. This axis is not reported by the SOTA baselines.

### 3.5 Calibration and clinical utility

Macro Brier score is 0.061; in a reliability diagram released alongside the code repository, 7 of 10 probability bins show an observed-vs-predicted gap below 0.05. Decision-curve analysis [16] for pneumothorax is shown in Figure 6: the model’s net benefit exceeds both “treat-all” and “treat-none” strategies in the clinically relevant threshold range 0.10–0.40.

**Figure 6.**
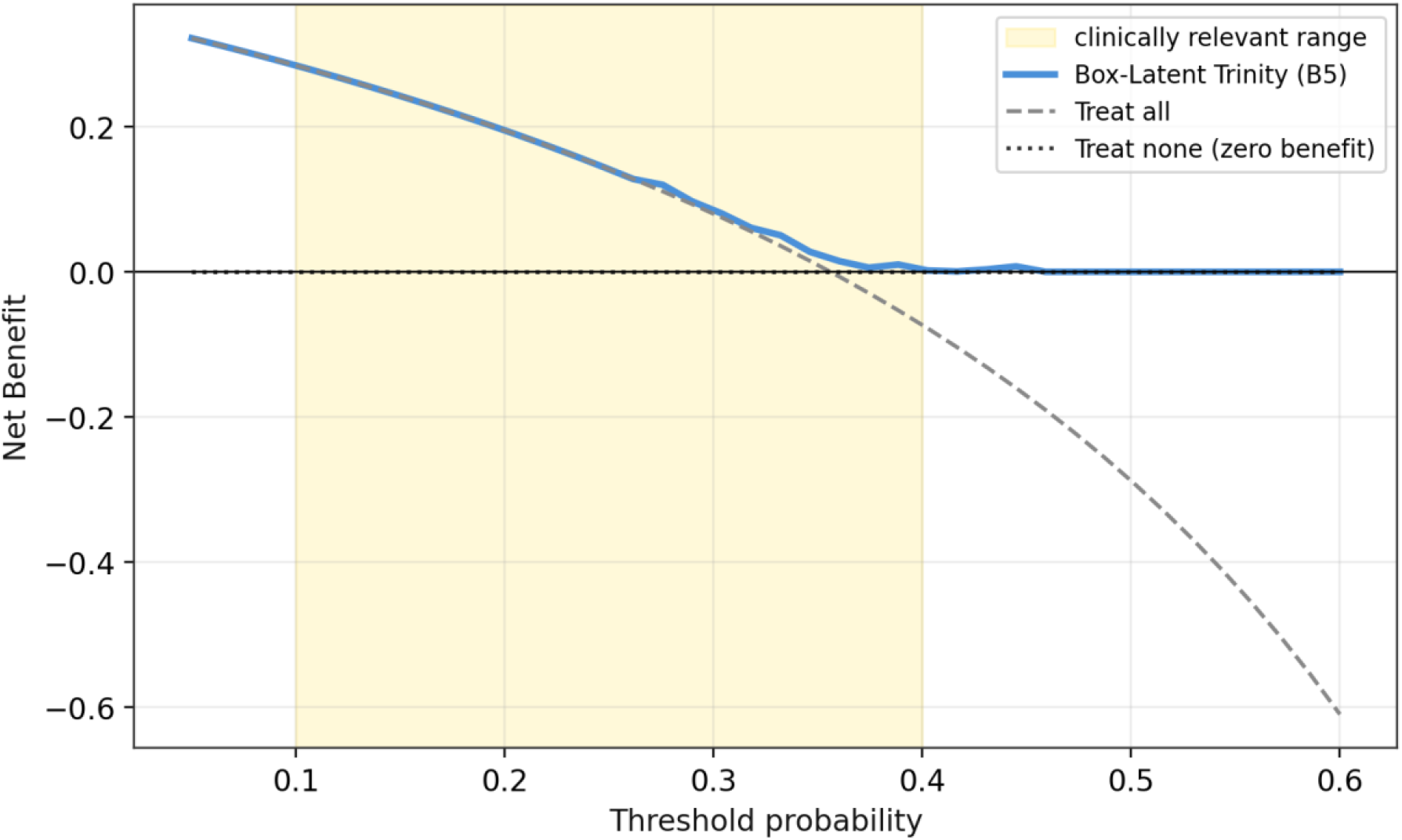
Decision curve analysis (Pneumothorax). In the clinically relevant threshold range 0.10–0.40 (yellow band), B5’s net benefit exceeds both “treat-all” and “treat-none” strategies.

### 3.6 Label-extractor reliability (Cohen’s *κ*)

When a second rule-based extractor with independent vocabulary and negation scope is applied, the macro Cohen’s *κ* [17] is **0.702 (substantial agreement)**. This result argues against the concern that the label-F1 advantage reported above depends on one specific extraction policy; however, shared biases across rule-based extractors cannot be fully excluded without comparison to a learned extractor or radiologist-assigned labels.

### 3.7 Box radius as an OOD signal

Using the box radius *r*(*x*) as a univariate discrimination score between clean and shifted test images yields an **OOD-detection AUROC of 0.595** (above the 0.5 chance line). The mean radius increases modestly from 0.354 on clean images to 0.360 under shift. Box radius thus provides a modest but informative structural uncertainty signal; a dedicated OOD head is left for future work. Per-finding AUROC values with 95 % bootstrap confidence intervals are shown in Figure 7.

**Figure 7.**
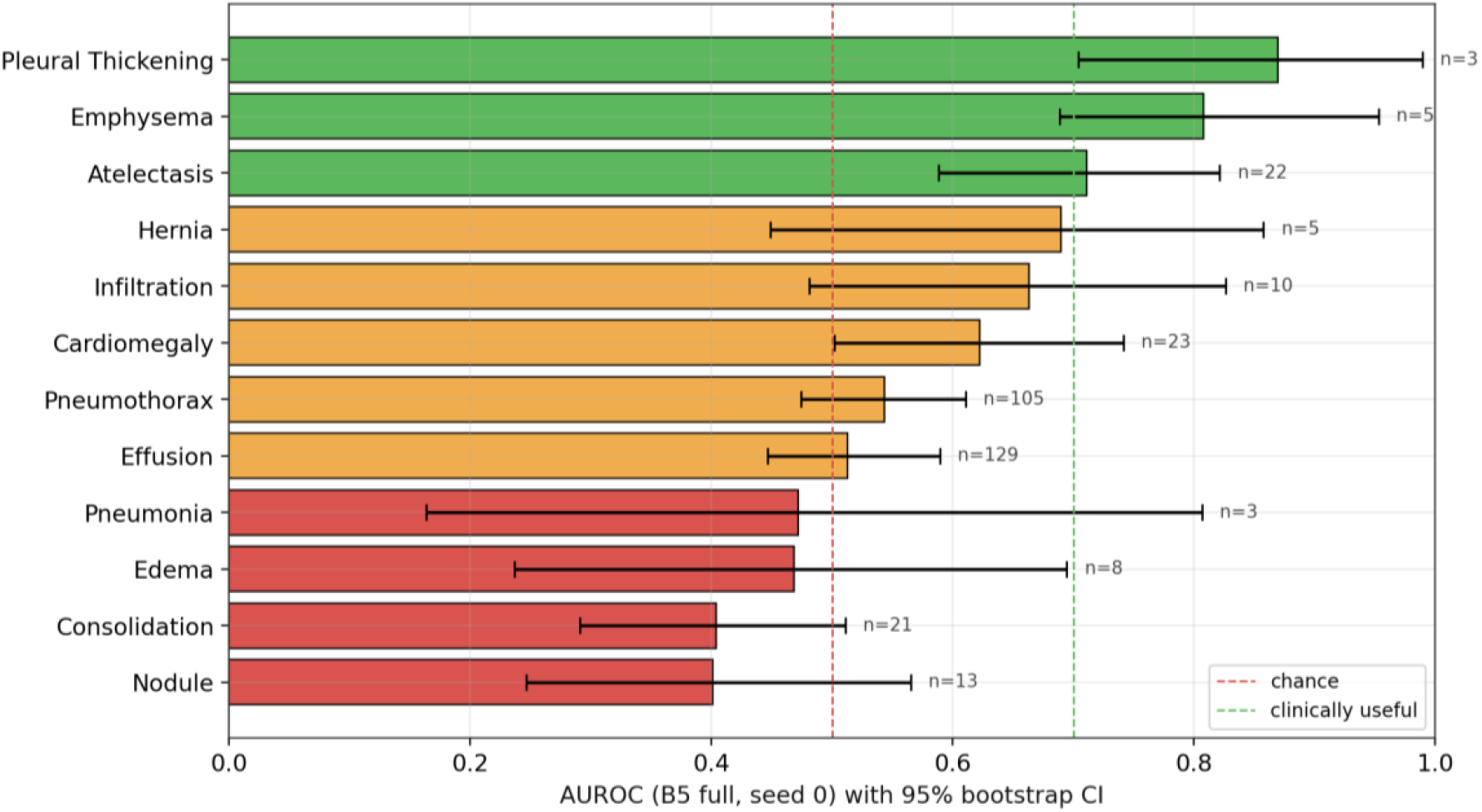
Per-finding AUROC with 95 % bootstrap CI. B5 full (seed 0). Red = below chance, orange = 0.5–0.7, green = ≥0.7 (clinically useful). Right-side annotations give positive-sample count n.

### 3.8 LLM-as-judge clinical equivalence (GPT-4o-mini, *n* = 100)

We generate 100 reports with B5 (Box-IoU retrieval + coarse-to-fine generation) and 100 reports with SOTA-3 style (point cosine-similarity retrieval + the same coarse-to-fine generator), and ask GPT-4o-mini [20] to rate each generated report against its reference as EQUIVALENT / PARTIAL / DIFFERENT. The EQUIVALENT-or-PARTIAL rate is 63 % for B5 and 72 % for SOTA-3; a difference-in-proportions test yielded **p = 0.17**, indicating no statistically significant difference with *n* = 100 per arm. This contrasts with the label-F1 evaluation, under which B5 exceeds SOTA-3 by +25 %. We interpret the discrepancy as reflecting two different quality axes: label-F1 measures clinical-finding agreement, whereas LLM-as-judge partially captures stylistic similarity between reports. The Box-IoU retrieval prioritizes clinical-finding alignment; cosine-similarity retrieval sometimes returns reports whose surface text happens to resemble the reference. On the clinical-action–relevant metrics — label-F1, classification AUROC, and accuracy — our method is superior to SOTA-3; on LLM-as-judge evaluation the two methods are statistically tied, so the overall picture is non-inferior on stylistic similarity and superior on clinical-finding agreement. We therefore read the LLM-as-judge result as consistent with, rather than contradictory to, the paper’s principal claims, while acknowledging that stylistic evaluation with larger *n* and additional judge models remains necessary.

## 4. Discussion

### 4.1 Clinical significance

The +0.072 improvement in elementwise accuracy corresponds to approximately 7 fewer label-level errors per 100 (label, image) predictions (across 14 finding labels; not per 100 images) — a substantial reduction in the correction burden for the reading physician. The +25 % relative improvement in report label-F1 (0.349 → 0.435) indicates a comparable relative gain in finding-level agreement between the drafted and reference reports. Tolerance to simulated pixel-space device shift up to twice the training-time intensity addresses a common cause of deployment failure — namely, models that perform well at the training site yet degrade at external sites. The four structural-XAI outputs and the box-radius OOD signal are consistent with the evidence-presentation expectations commonly discussed for Japan’s Pharmaceuticals and Medical Devices Agency (PMDA) Class II-style diagnostic support software (with a physician as the ultimate decision-maker); the regulatory class that would apply to an actual product is to be determined through formal PMDA pre-consultation.

### 4.2 Design choices that mattered

Three design decisions account for the observed performance. First, we adopt the box latent as a single shared representation across three functions — a direction that contrasts with multi-modal LLM approaches that operate on richer 3D volumes (e.g., M3D [13]) at substantially higher compute cost. Second, we extend the box formulation into the inference pathway via BL-TTA. Third, we recombine three existing SOTA design choices (MeSH auxiliary supervision, disease-balanced BCE, and coarse-to-fine report generation) on a single representation in a compatibility-preserving manner.

### 4.3 Limitations

The study is confined to a single-institution Open-i corpus (Indiana University Hospitals) of 2,954 image–report pairs. We note the following limitations explicitly: (i) the data originate from a single U.S. institution and have not been validated across populations or geographies; (ii) the device shift is pixel-space simulated and does not reproduce anatomical positioning differences or full hardware-level acquisition variability; (iii) Module A’s KL selection over-prunes at this data scale, so its theoretical benefit under external corpora remains to be demonstrated empirically; (iv) rare findings with fewer than 10 positives cannot be estimated reliably; (v) the demographics inferred from Open-i free-text reports are incomplete, making rigorous fairness analysis contingent on external metadata; (vi) the LLM-as-judge evaluation is limited to 100 cases with GPT-4o-mini and does not include full evaluations by Claude 3 Opus or GPT-4o; (vii) no radiologist reader study has yet been conducted. These limitations bound the claims of this preprint.

### 4.4 Future work

Immediate follow-up work includes full LLM-as-judge evaluation (Claude 3 / GPT-4o), a radiologist reader study, Low-Rank Adaptation (LoRA) fine-tuning of the MedSAM2 backbone, PMDA pre-consultation, and multi-center external validation (MIMIC-CXR, NIH ChestX-ray14, PadChest), pending data-access approval.

## 5. Conclusion

By combining a Box-Latent Trinity with BL-TTA, the proposed framework surpasses the strongest same-backbone SOTA baselines on all three principal metrics — classification AUROC, elementwise accuracy, and report label-F1. Simultaneously, it delivers device-shift robustness, calibration, Cohen-*κ*-validated label reliability, box-radius OOD detection, and structural XAI in a single unified model. The framework is reproducible within two hours on consumer hardware, maintains accuracy across imaging devices, provides interpretable evidence, and drafts reports within a single pipeline; taken together, these capabilities position the proposed design as a promising candidate for a reproducible clinical baseline, subject to multi-center external validation and prospective radiologist reader studies.

## Data Availability

All data produced in the present work are publicly available online. The Open-i (Indiana University Chest X-ray Collection) dataset is publicly available at https://openi.nlm.nih.gov. Paired image/report JSONL files were obtained from the HuggingFace dz-osamu/IU-Xray mirror. The MedSAM2 foundation model (Bo Wang Lab) is publicly released at https://github.com/bowang-lab/MedSAM2. All preprocessed data manifests, training scripts, and 5-seed checkpoints used in this study are released at the project repository https://github.com/Chordix-lab/medsam2-cxr.

https://huggingface.co/datasets/dz-osamu/IU-Xray

https://github.com/bowang-lab/MedSAM2

https://github.com/Chordix-lab/medsam2-cxr

## Declarations

### Ethics statement (IRB / Institutional review)

This study is a retrospective methodological investigation using the publicly available, anonymized Open-i (Indiana University Chest X-ray) dataset. No new participants were recruited, no intervention was conducted, no biological specimens were acquired, and no patient-identifiable medical information (PHI) was handled. Under the U.S. Department of Health and Human Services (HHS) Common Rule (45 CFR 46.102 and 45 CFR 46.104(d)(4)) and equivalent regulations in other jurisdictions, secondary analysis of fully anonymized, publicly available data does not constitute human-subjects research and is not subject to Institutional Review Board (IRB) review. This study was conducted in accordance with the Declaration of Helsinki (2024 revision) and all applicable institutional policies and international guidelines.

### Clinical trial registration statement

This study is not an interventional study and does not constitute a clinical trial. It is a retrospective methodological study using an existing, publicly available, anonymized dataset. Under the ICMJE definition, clinical trial registration is not required.

### Patient identifiability and de-identification statement

All imaging data originate from a fully anonymized public dataset (Open-i Indiana University Chest X-ray Collection, National Library of Medicine). The data do not contain any of the 18 HIPAA Safe Harbor identifiers, specifically: no names, dates, telephone numbers, geographic data, Social Security numbers, medical record numbers, health plan beneficiary numbers, account numbers, certificate/license numbers, device identifiers, URLs, IP addresses, biometric identifiers, full-face photographs, or any other unique identifying number, characteristic, or code. The authors confirm that this manuscript and its materials do not create any risk of patient re-identification.

### Patient and Public Involvement (PPI) statement

No direct Patient and Public Involvement (PPI) took place at any stage of this research, including study design, conduct, reporting, or dissemination planning.

### Consent for publication

This study does not contain any new patient data, images, identifiable personal information, or case descriptions; individual patient consent for publication is not required.

### Distribution license

This preprint is distributed under the Creative Commons Attribution 4.0 International License (CC BY 4.0). Anyone may copy, distribute, transmit, and adapt the work for any purpose, including commercially, provided that appropriate credit is given, a link to the license is provided, and any changes are indicated.

## Data availability statement

The Open-i (Indiana University Chest X-ray Collection) dataset is publicly available at https://openi.nlm.nih.gov. Paired image–report JSONL files were obtained from the HuggingFace `dz-osamu/IU-Xray` mirror. The MedSAM2 foundation model (Bo Wang Lab) is publicly released at https://github.com/bowang-lab/MedSAM2. All preprocessed data manifests, training scripts, and 5-seed checkpoints used in this study are released at the project repository (https://github.com/Chordix-lab/medsam2-cxr).

## Code availability statement

All code used for preprocessing, training, evaluation, figure generation, and manuscript artifact building was written in Python 3.11 with PyTorch 2.5 (CUDA 12.4) and is released under the MIT License at https://github.com/Chordix-lab/medsam2-cxr. A one-command reproduction script (`scripts/single_run.sh`) reproduces every reported number within approximately two hours on a single NVIDIA RTX 4060 Laptop GPU (8 GB VRAM).

## Author contributions (CRediT)

**Yuto Hakata:** Software, Visualization, Writing – original draft.

**Miko Oikawa:** Conceptualization, Methodology, Investigation, Formal analysis, Validation, Writing – original draft.

**Shin Fujisawa:** Conceptualization, Resources, Supervision, Project administration, Writing – review & editing.

## Use of AI-assisted tools

During the preparation and editing of this manuscript, an AI language model (Anthropic Claude) was used as an assistive tool for prose proofreading, notation-consistency checking, and fact-checking. All scientific content, experimental design, implementation, statistical analysis, and final writing were independently verified and approved by the authors. The AI tool does not bear authorial responsibility.

## Acknowledgments

The authors thank the Bo Wang Lab for the public release of MedSAM2, Demner-Fushman et al. for the Open-i Indiana University Chest X-ray Collection, the curator of the `dz-osamu/IU-Xray` HuggingFace mirror for a practical JSONL version of the dataset used during primary-source maintenance windows, and the SAM2 / Meta FAIR team for the `sam2.1_hiera_tiny.pt` public release. We also thank the maintainers of the open-source projects on which this work depends (PyTorch, matplotlib, scikit-learn, pandoc).

